# Forecasting COVID-19 Number of Cases by Implementing ARIMA and SARIMA with Grid Search in United States

**DOI:** 10.1101/2021.05.29.21258041

**Authors:** Saina Abolmaali, Samira Shirzaei

## Abstract

COVID-19 has surged in the United States since January 2020. Since then, social distancing and lockdown have helped many people to avoid infectious diseases. However, this did not help the upswing of the number of cases after the lockdown was finished. Modeling the infectious disease can help the health care providers and governors to plan ahead for obtain the needed resources. In this manner, precise short-term determining of the number of cases can be imperative to the healthcare system. Many models have been used since the pandemic has started. In this paper we will compare couple of time series models like Simple Moving Average, Exponentially Weighted Moving Average, Holt-Winters Double Exponential Smoothing Additive, ARIMA, and SARIMA. Two models that have been used to predict the number of cases are ARIMA and SARIMA. A grid search has been implemented to select the best combination of the parameters for both models. Results show that in the case of modeling, the Holt-Winters Double Exponential model outperforms Exponentially Weighted Moving Average and Simple Moving Average while forecasting ARIMA outperforms SARIMA.

## 1 Introduction

As of May 10 total number of 32,543,257 people in the United States are infected with COVID-19 [1]. Due to the mysterious nature of the virus, quarantine was the first and continuous response used to prevent the spread of the disease. During the pandemics, several strategies were adopted toward controlling the spread of the disease. Statistical and mathematical modeling have been helping politicians and healthcare governors control and prepare for the outbreak and adopt various strategies. Data analytic have been used in various researches such as medical and finance[2]. Time-series techniques are prevalent in modeling and predictions of the data series indexed with time. One study have used Linear regression, SIR model and logistic regression to predict the number of infected individuals for four countries.[3]. A study performed by Chen et al. used the ARIMA model to forecast property crime in china. They have shown that the ARIMA model prediction is very accurate [4]. SARIMA has been used by Szeto et al. to model and forecast traffic. The results showed only a ten percent error in the forecast [5]. Time-series analyses have been used in many publications to predict and forecast the number of infected by the infectious disease. For example, in a study, Lai modeled the number of individuals infected by SARS through the Box-Jenkins model and ARIMA [6]. Martinez in his paper has used SARIMA to model the incidence of dengue. The results indicate that the SARIMS model can help disease control predict the number of cases precisely [7]. In another work, Roy et al. used the ARIMA model to represent the COVID-19 spread, and to evaluate their model, they have calculated RMSE and MAE[8]. Tandon et al. predicted the rise in the number of cases for a short time using ARIMA time series[9]. Koyuncu et al. have used SARIMA to investigate how COVID-19 has affected maritime by forecasting RWI/ISL [10]. In another study, Ceylan has used ARIMA and SARIMA models to investigate the prevalence of COVID-19 in three countries of Italy, France, and Spain [11]. ARIMA Models have been used in many papers to predict the overview of the infectious diseases [12], [13], [14], [15], [16], [17], [18]. In this paper, we are exploring five time-series models. We use SMA, EWMA, and Holt Winter Double Exponential to describe the data and model the prevalence of the confirmed cases and deceased cases. Afterward, we use ARIMA and SARIMA to model and predict the number of infected individuals. All the analyses are performed on four states of the United States. States are selected from 4 different geographic locations to describe the pandemic distribution perfectly. Selected states are Alabama, Massachusetts, California, and Washington. All of the analyses are performed for both the number of confirmed cases and the number of deceased cases. For evaluation of the models’ performance and predictions, RMSE (Root Mean Square Error) and MSE (Mean Square Error) indicate the precision of each prediction. As a result, the performance of the three models, SARIMA and ARIMA, are compared based on the modeling and prediction. As the number of reported cases is the number of positive tested individuals, the data might not be accurate. There are many infected individuals that they do not take a test or they would not use medical treatment for the disease.

## 2 Method

### 2.1 Data

To perform further analysis on the United States data set, We are using the John Hopkins data repository of COVID-19, the most reliable source of data on COVID-19 [19]. Time series data provided by John Hopkins GitHub repository consists of the daily number of confirmed cases and related states. Due to the high number of data and the fact that modeling all the states is away from the scope of the paper, we are just representing the model and forecasting for four states, including Alabama, Washington, California, and Massachusetts.

### 2.2 Simple Moving Average

As mentioned before, we apply the Simple Moving Average technique to model the infected and deceased cases. The simple moving average is calculated through a total of the recent number of cases and dividing by the total number of periods involved. The formula would be as follow:

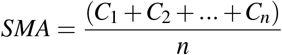

We model four states with two parameters of 10 days and 30 days with the simple moving average. We can see here in the case of confirmed cases, as the moving average parameter goes up, the model performance decreases, which is reasonable due to the nature of the simple moving average. Also, for short-term modeling, a simple moving average performs fine. In all eight figures, we can see a decrease in the number of cases for late March.

**Fig. 1.**
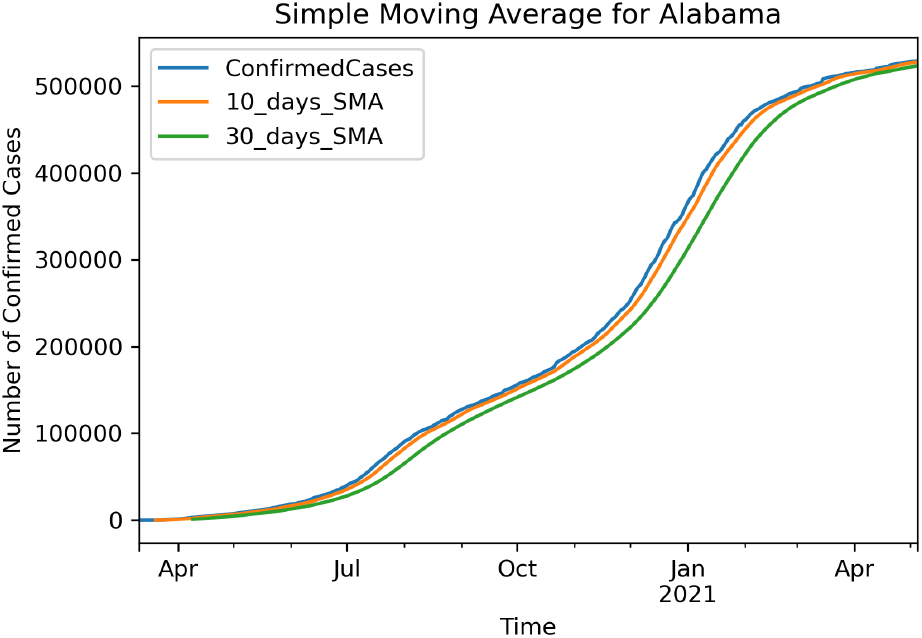
Simple Moving Average for Confirmed Cases Alabama

**Fig. 2.**
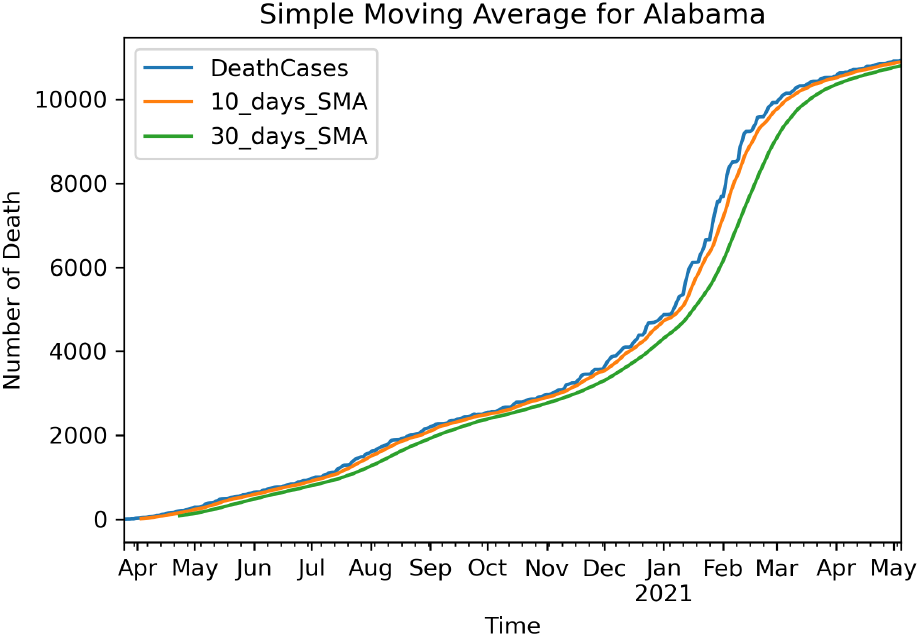
Simple Moving Average for Deceased Cases Alabama

### 2.3 Exponentially Weighted Moving Average 30 Days

The primary difference between an EWMA and an SMA is the sensitivity of each one to the data. The sensitivity to the data means SMA assigns uniform weight to all of the data, while EMA gives more weight to current data. The newest data will impact the moving average more, while older data has less impact on the average. The EWMA is calculated as follow:

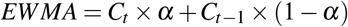

**Fig. 3.**
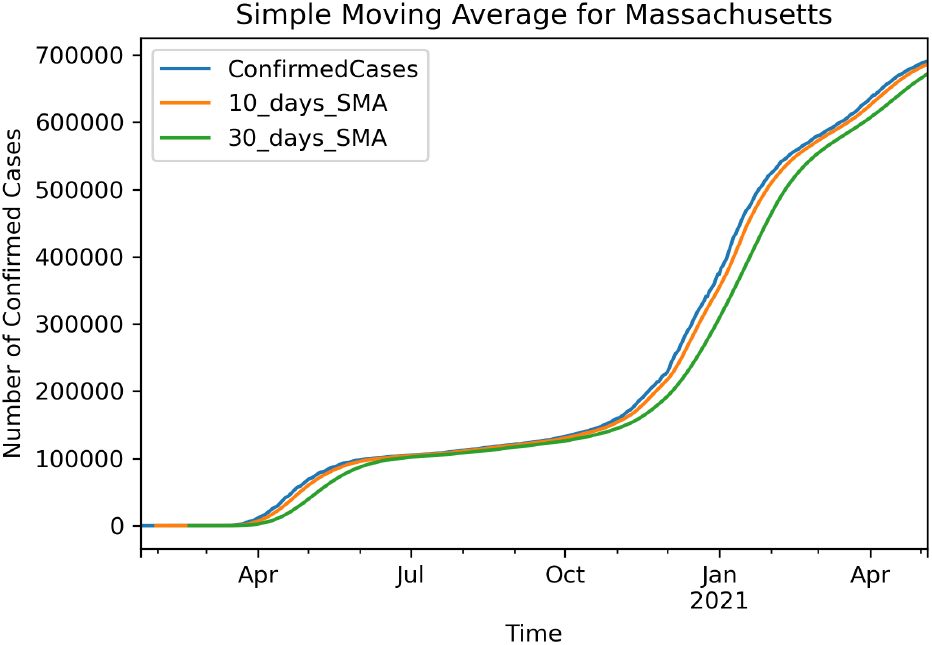
Simple Moving Average for Confirmed Cases Massachusetts

**Fig. 4.**
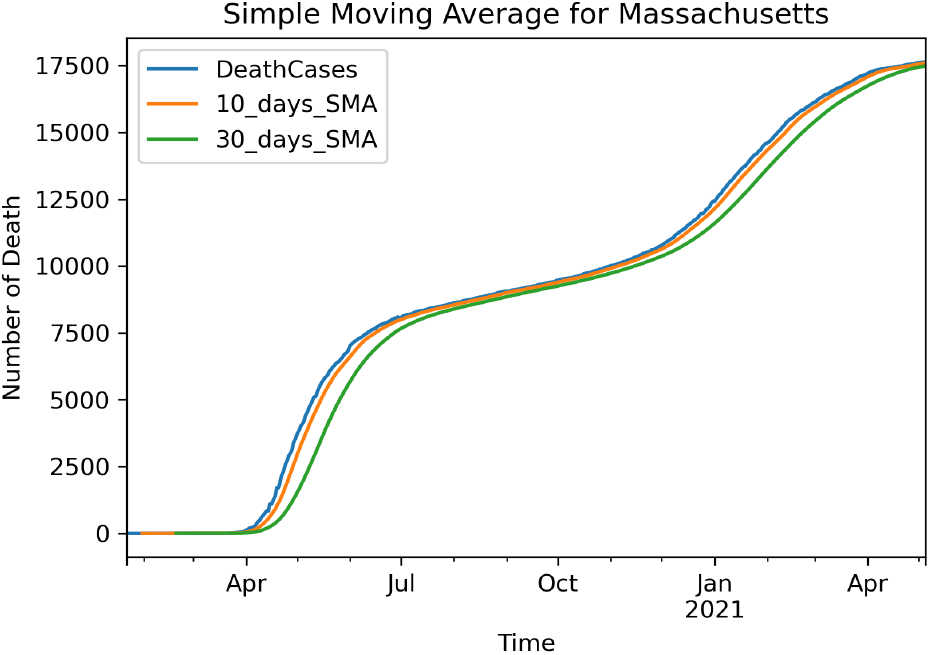
Simple Moving Average for Deceased Cases Massachusetts

**Fig. 5.**
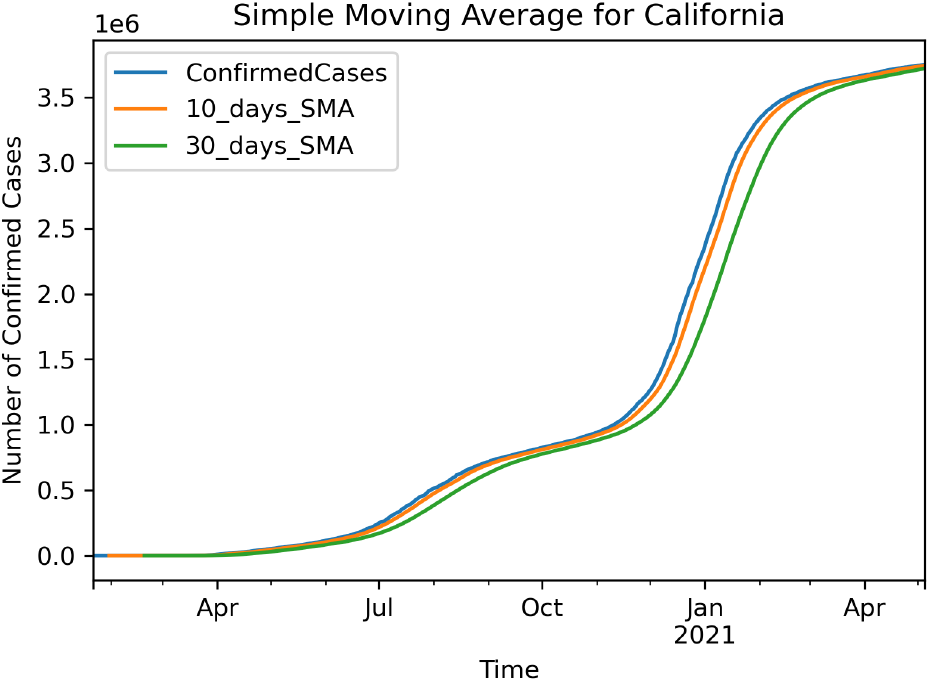
Simple Moving Average for Confirmed Cases California

We use 30 days of the recent data to calculate EWMA. The following plots show the EWMA on the data of four states with a parameter of 30 days. To compare these models together, we can see if the SMA parameter goes higher, it can not represent the model well enough, and EWMA might be a better representation of the data in this case.

**Fig. 6.**
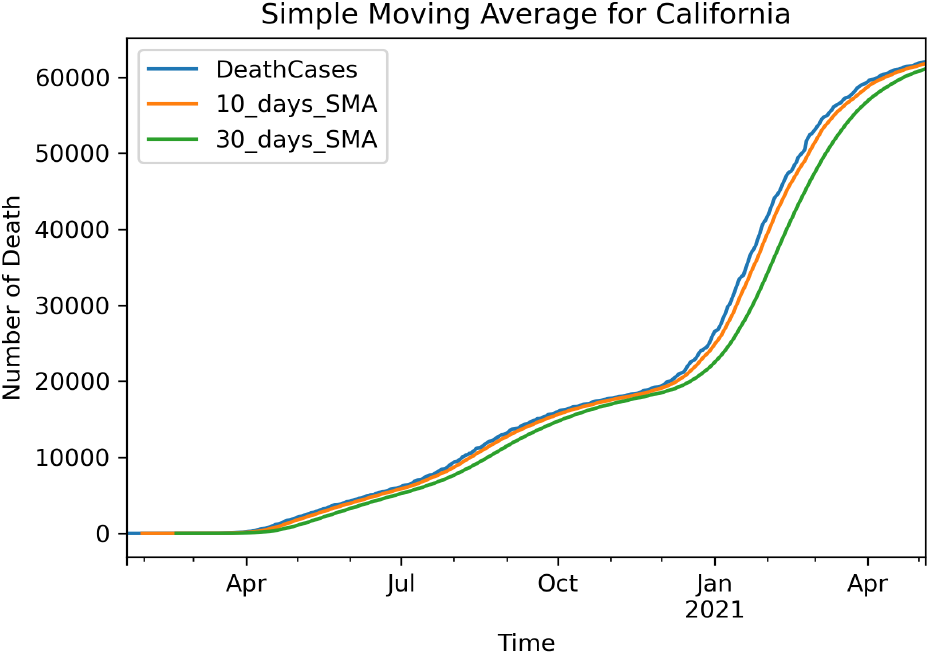
Simple Moving Average for Deceased Cases California

**Fig. 7.**
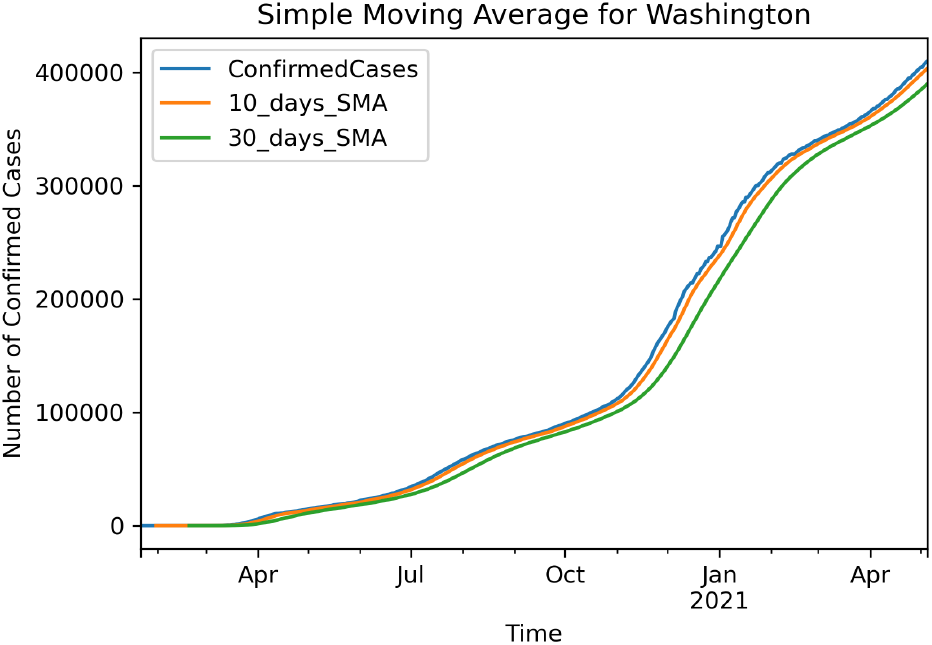
Simple Moving Average for Confirmed Cases Washington

**Fig. 8.**
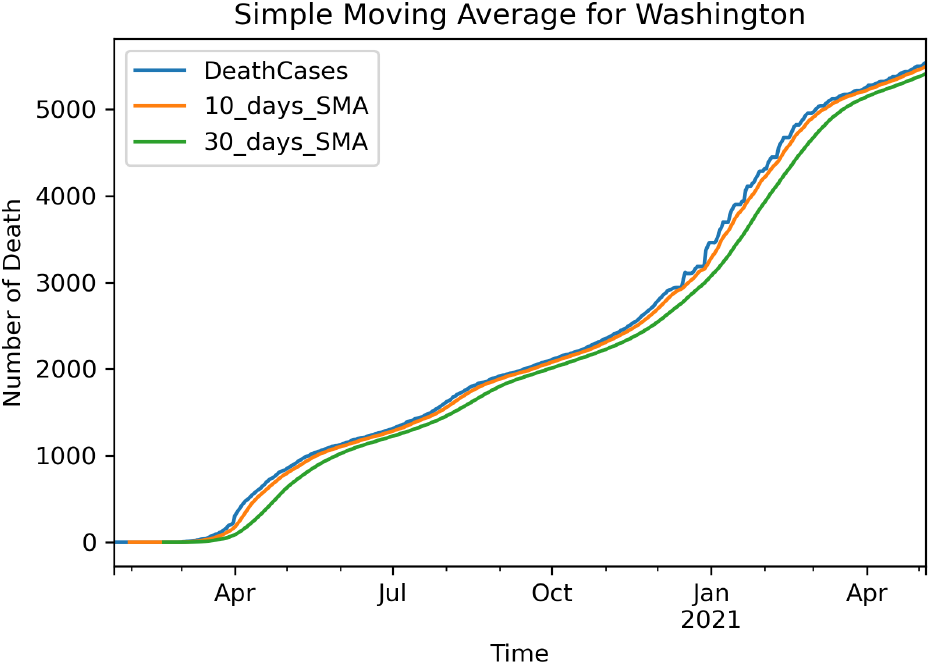
Simple Moving Average for Deceased Cases Washington

**Fig. 9.**
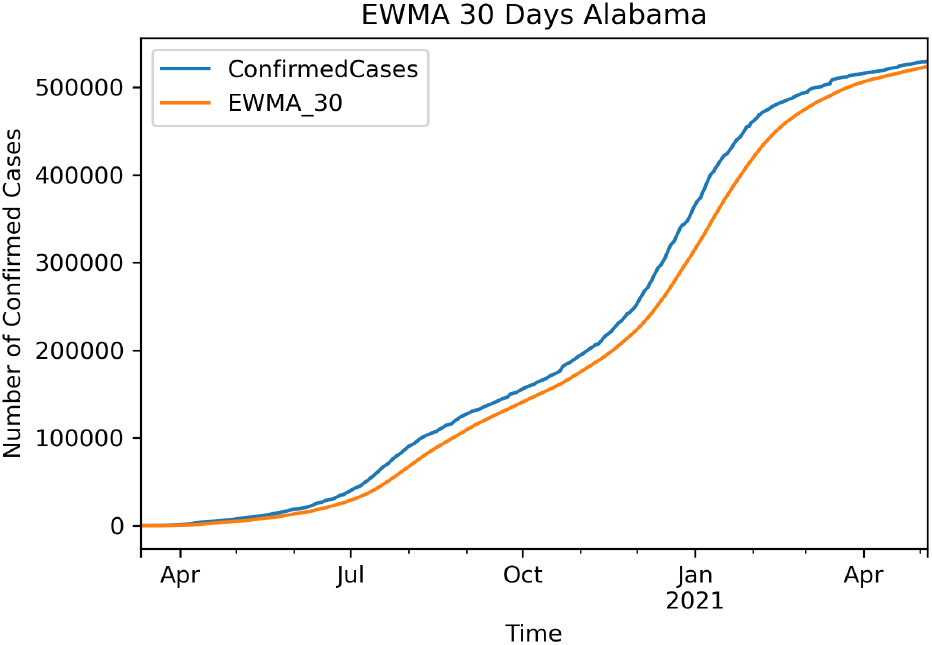
EWMA for confirmed Cases Alabama

**Fig. 10.**
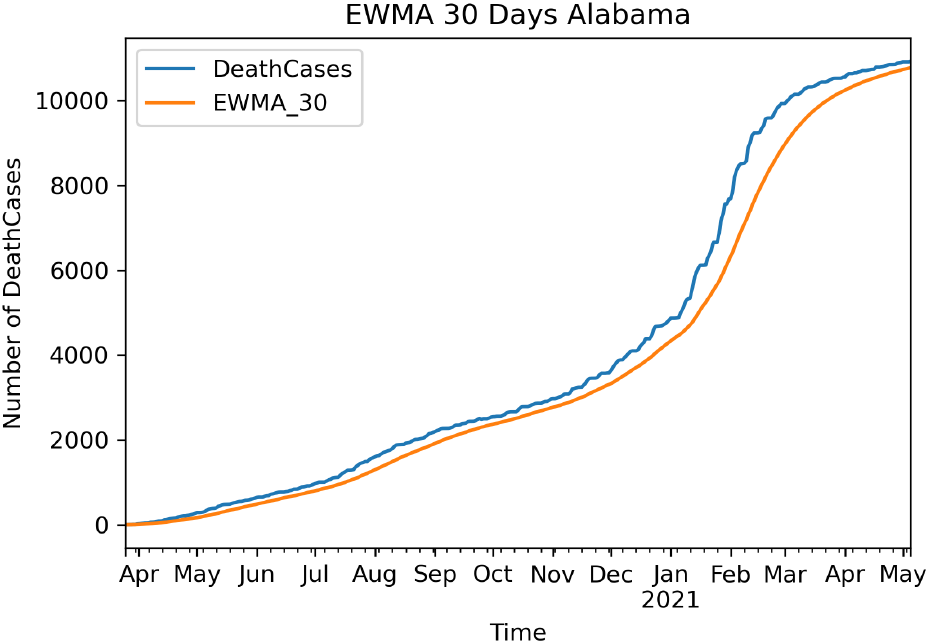
EWMA for deceased Cases Alabama

**Fig. 11.**
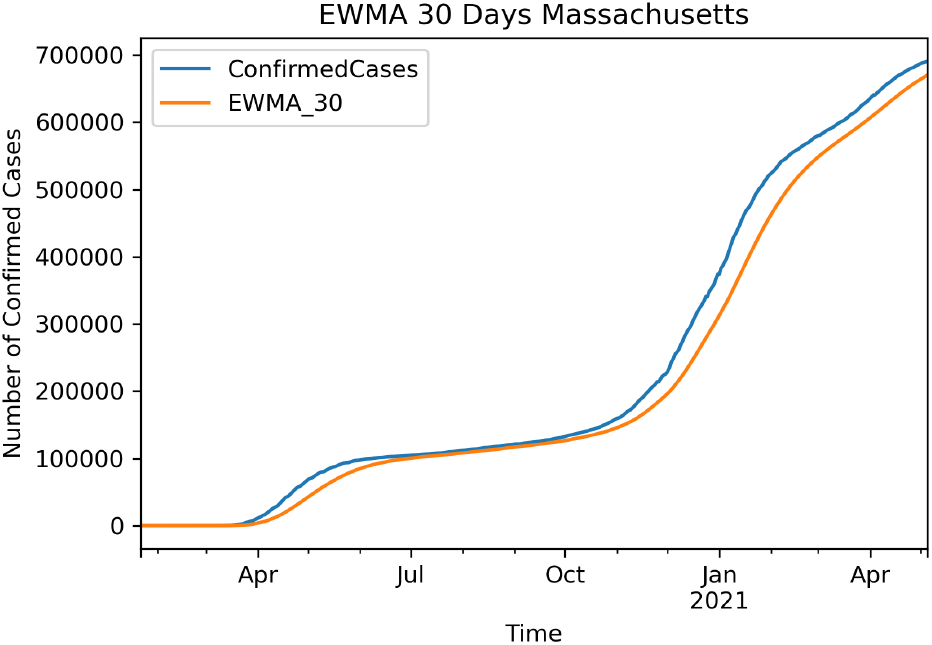
EWMA for confirmed Cases Massachusetts

### 2.4 Holt Winters Double Exponential Smoothing Additive

Exponential smoothing is a methodology for persistently reexamining a forecast in the light of later experience [20]. The Holt–Winters’ model consists of two variants: the additive and the multiplicative. The additive technique is suitable when the occasional varieties are generally steady through the series. The multiplicative technique is preferred when the seasonal variations are changing proportionally to the level of the series. Whether the occasional variety is viewed as independent of the level of the neighborhood mean or as being relative to it [21]. Winter argues that some other factors can influence seasonal estimates. The paper mentions that seasonal factors will rise to fill the lack of train [22]. Double Exponential models are used when the data has a trend. The Holt-Winter seasonal method contains three smoothing equations. The first equation is to update the level with parameter *α*, the second one is to update the trend with parameter *β*, and the third one is to update the seasonality with parameter *γ*. We use the model as the following:

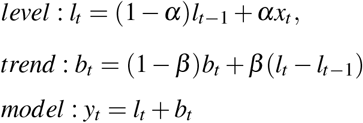

**Fig. 12.**
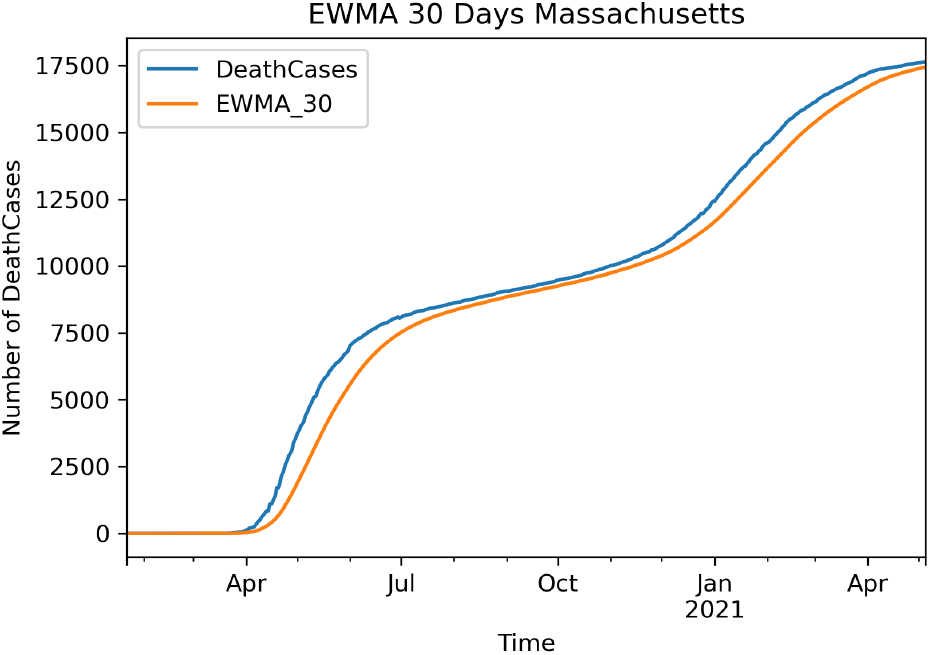
EWMA for deceased Cases Massachusetts

**Fig. 13.**
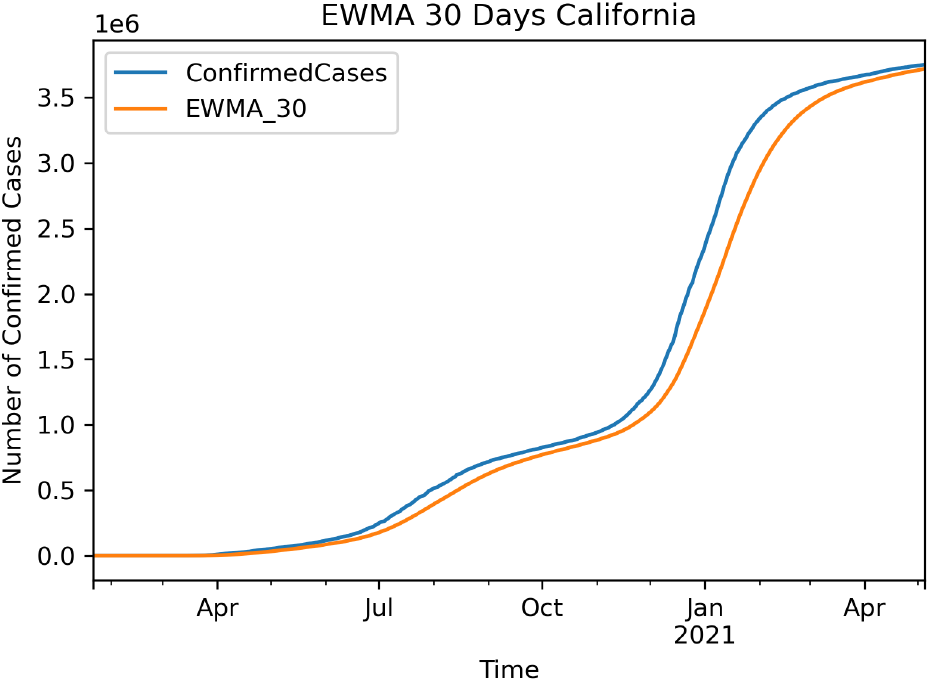
EWMA for confirmed Cases California

**Fig. 14.**
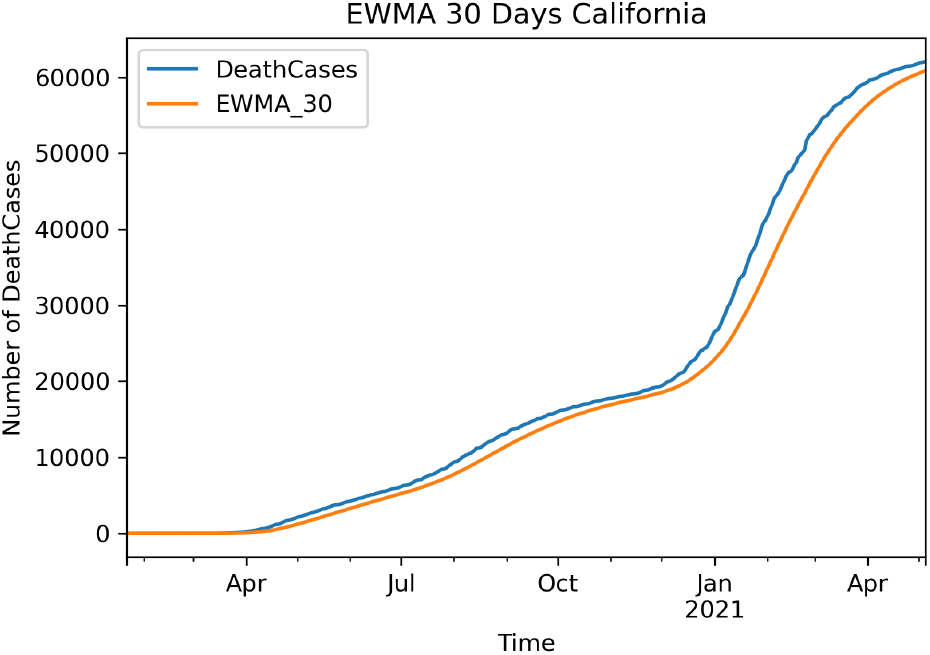
EWMA for deceased Cases California

**Fig. 15.**
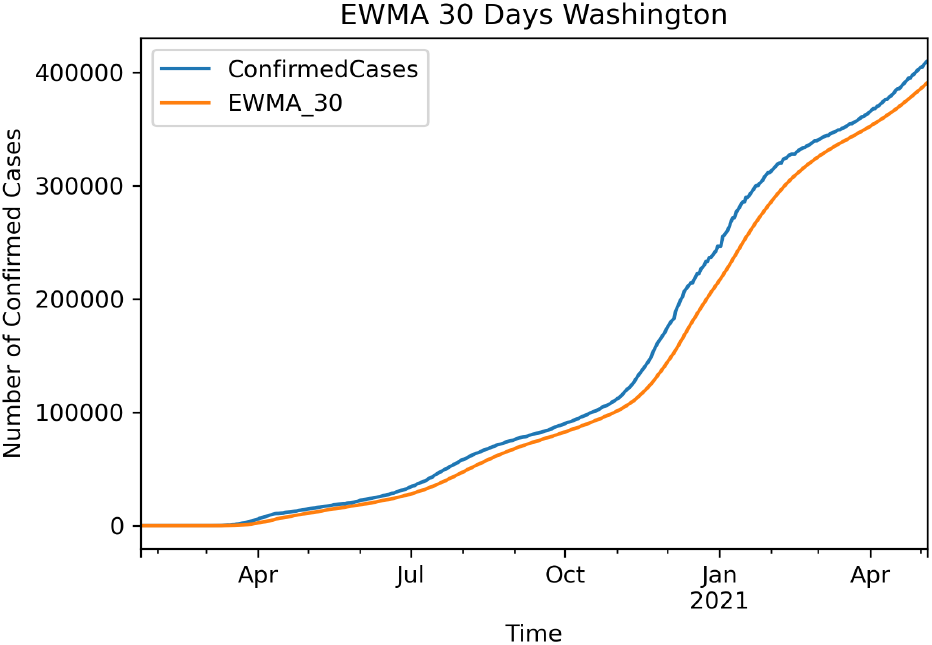
EWMA for confirmed Cases Washington

**Fig. 16.**
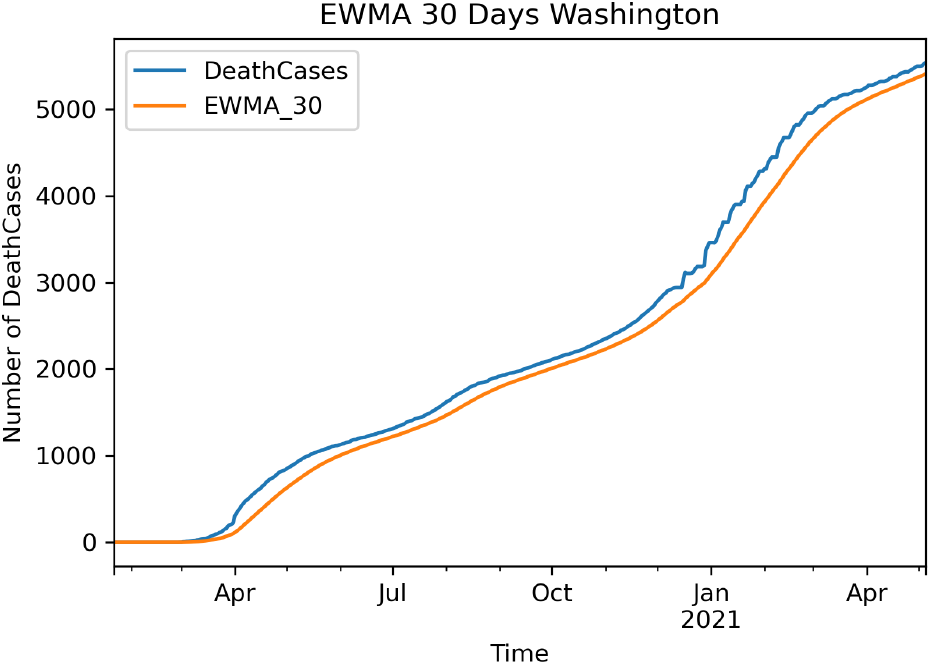
EWMA for deceased Cases Washington

Note that we use Holt Winter Double Exponential Smoothing just for modeling, not for prediction. Results show that Holt-Winter Double Exponential Smoothing can be a great representative of the models with trend and seasonality. Although due to the seasonal variations, which are roughly constant through the series, it is evident that the additive model would perform better rather than the multiplicative model, we still apply both models. Based on the RMSE, we can conclude that the additive model is outperforming the multiplicative model on COVID-19 data. For both models, span = 30 days and 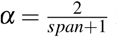 is selected. In the following figures, we can see HW model is outperforming EWMA30.

**Fig. 17.**
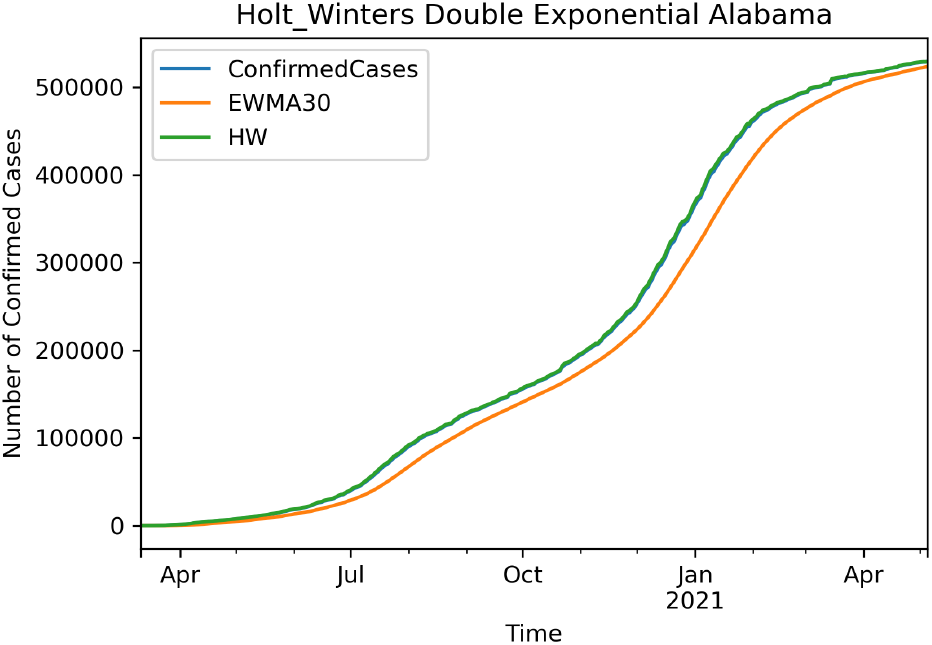
H-W Double Exponential for confirmed Cases Alabama

**Fig. 18.**
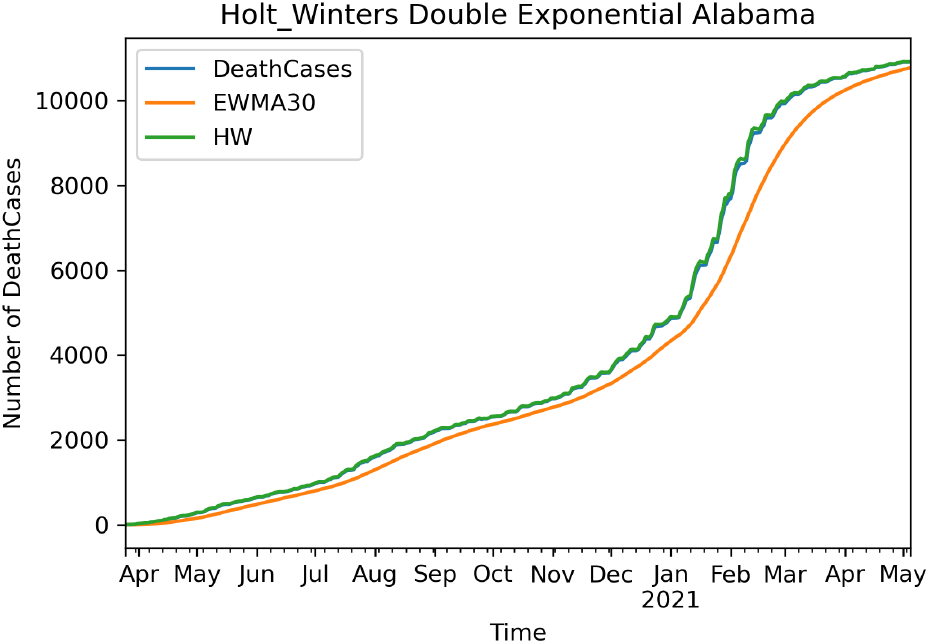
H-W Double Exponential for deceased Cases Alabama

### 2.5 ARIMA Model

ARIMA model or, ‘Auto-Regressive Integrated Moving Average’ model is a class of models that explains autocorrelations in the data. Box et al. first introduced the ARIMA model in the 1970s [23]. Using the data and the difference between time series values, ARIMA models can predict future values of the time series. ARIMA models are used in situations where data is non-stationarity. Therefore, the non-stationarity can be removed by applying differencing one or a couple of times. AR stands for Auto Regression, I as Integrated, and MA for Moving Average, which are three parameters of the ARIMA models. P stands for the order of the AR model, d the degree of difference, and q as the order of the MA model. We know that ARIMA(p,q,d) is defined as:

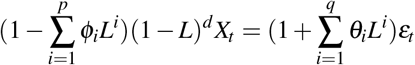

**Fig. 19.**
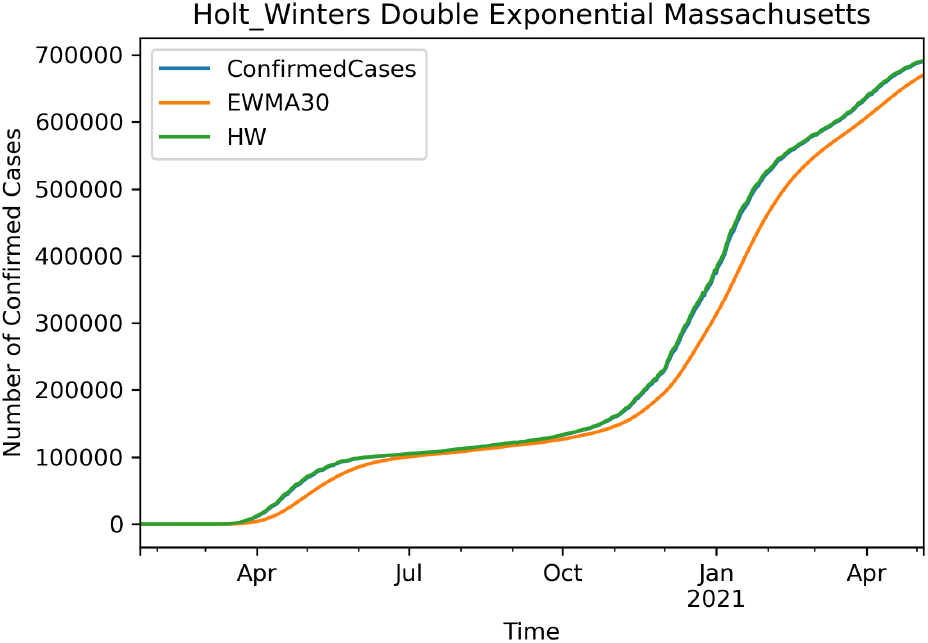
H-W Double Exponential for confirmed Cases Massachusetts

**Fig. 20.**
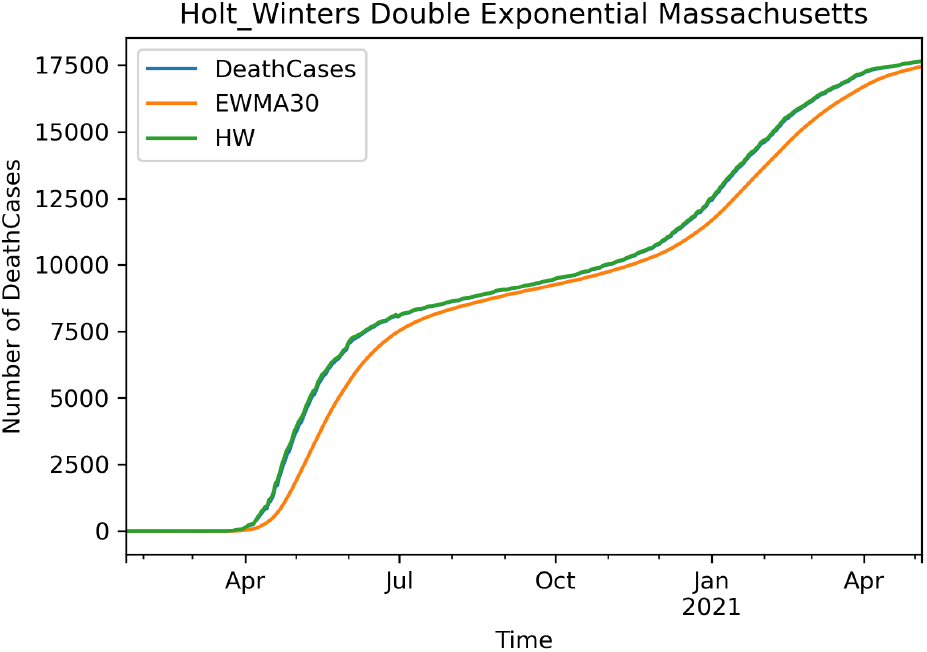
H-W Double Exponential for deceased Cases Massachusetts

**Fig. 21.**
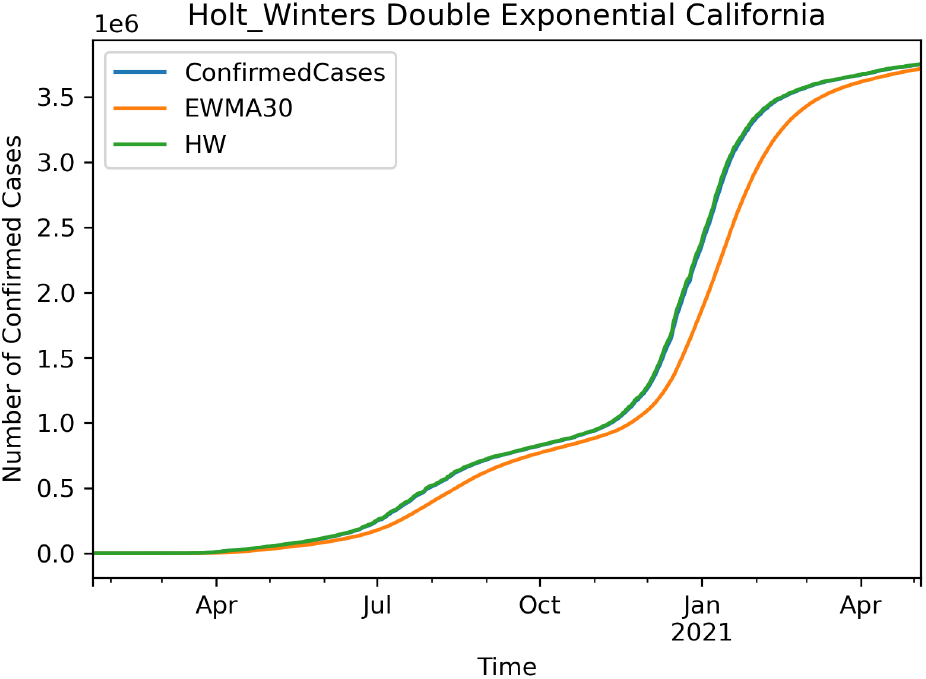
H-W Double Exponential for confirmed Cases California

**Fig. 22.**
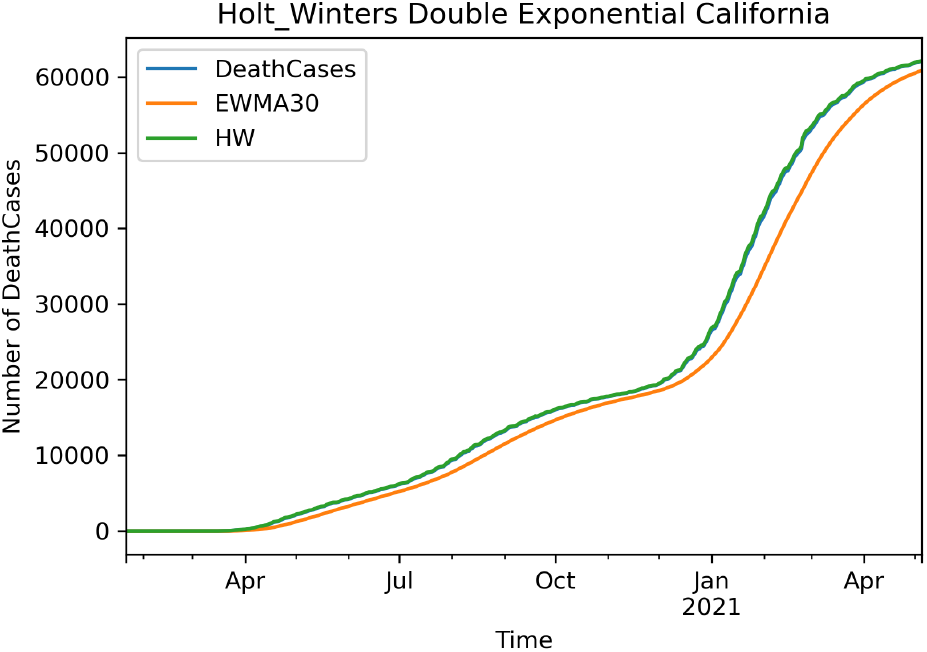
H-W Double Exponential for deceased Cases California

**Fig. 23.**
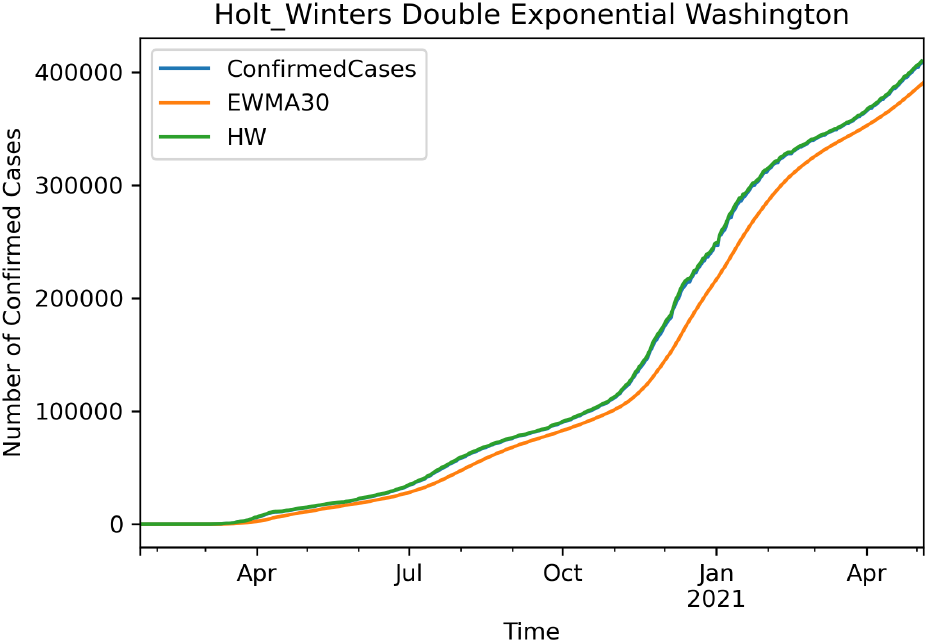
H-W Double Exponential for confirmed Cases Washington

**Fig. 24.**
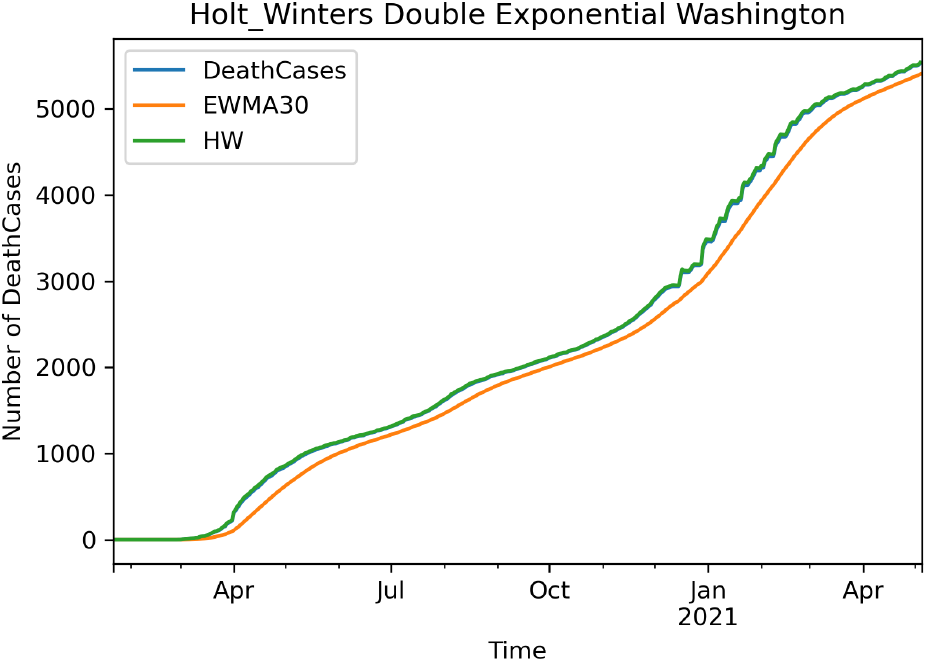
H-W Double Exponential for deceased Cases Washington

Where L is the lag operator. Here p and q represent how high of an order we should do for each of those components. We see p on the left-hand side for AR and q on the right-hand side for MA. Then we difference our data with the lag operator. In the ARIMA model, choosing the parameters by the programmer would not be the best choice since there might be human errors in reading the autocorrelation data, thus it would be better to use a grid search. To select the differencing terms, we used a test of stationary, augmented Dickey-Fuller test, and seasonality, the Canova-Hansen test for seasonal models. The best combination of p, d, and q has been chosen based on the AIC. RMSE (Root Mean Square Error) and MSE (Mean Square Error) have been used in this study to distinguish the ARIMA model accuracy for different states and compare different models together. We can see here that the model is performing differently for different states. We set the model as an iterative model to test different combinations of p,d, and q. This grid search provides the best combination to minimize the AIC. Each iteration provides an ARIMA model for the train data set. The grid search stores the lowest error for each iteration. Finally, the iteration stops after testing ten different models that would not perform better than the least error. Therefore, the parameters are selected, and the model is solid for forecasting. Forecasting is performed for 90 days. The model performs better for Alabama, and then the following states are California, Washington, and Massachusetts.

**Fig. 25.**
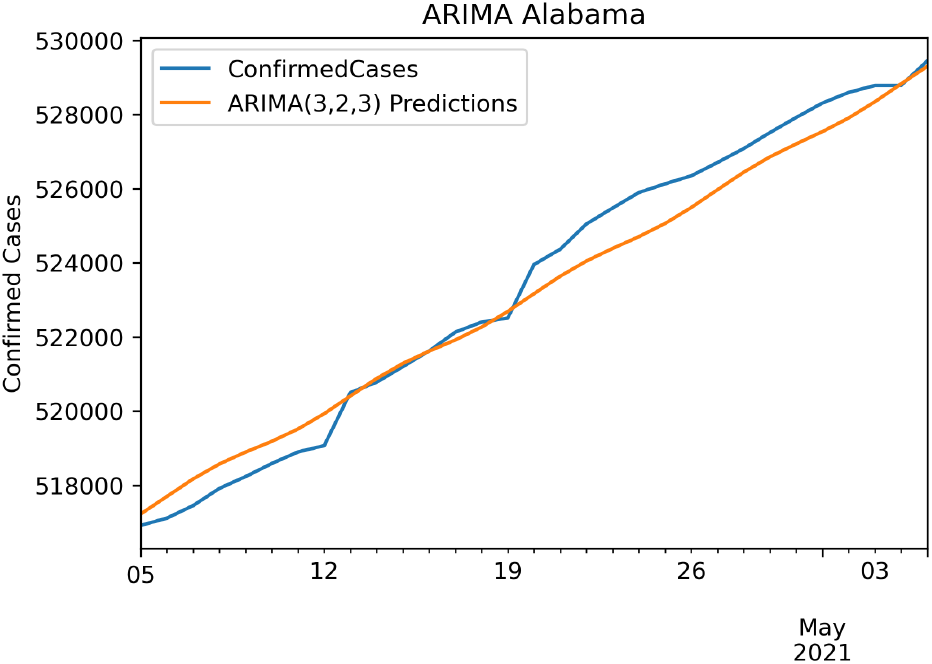
Alabama Confirmed ARIMA(3,2,3) RMSE: 653.2237608

### 2.6 SARIMA Model

SARIMA or Seasonal Autoregressive Integrated Moving Average specifically describes the seasonal models. Although the ARIMA model can support most of the time series, it cannot handle time series with seasonality. SARIMA model or Seasonal ARIMA are ARIMA models that contain a seasonal component in the model. ARIMA model omits seasonality by approaches like seasonal differencing. Here P represents seasonal regression, D represents differencing, Q represents moving average coefficient, and m represents the number of data points in each seasonal cycle. Again to determine the parameters, two approaches of ACF and PACF could be applied. In this paper, we have used the grid search algorithm mentioned in section 2.5. Each parameter we use for each state is stated in the figures. Again we use RMSE and MSE to measure the accuracy of the models. Forecasting is performed for 90 days, the same as the ARIMA model.

**Fig. 26.**
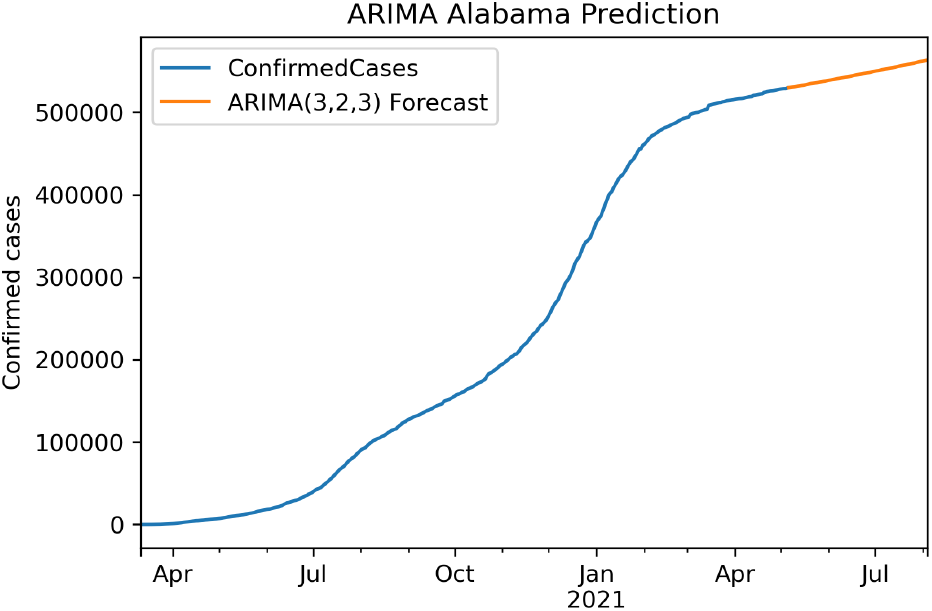
Alabama ARIMA Prediction

**Fig. 27.**
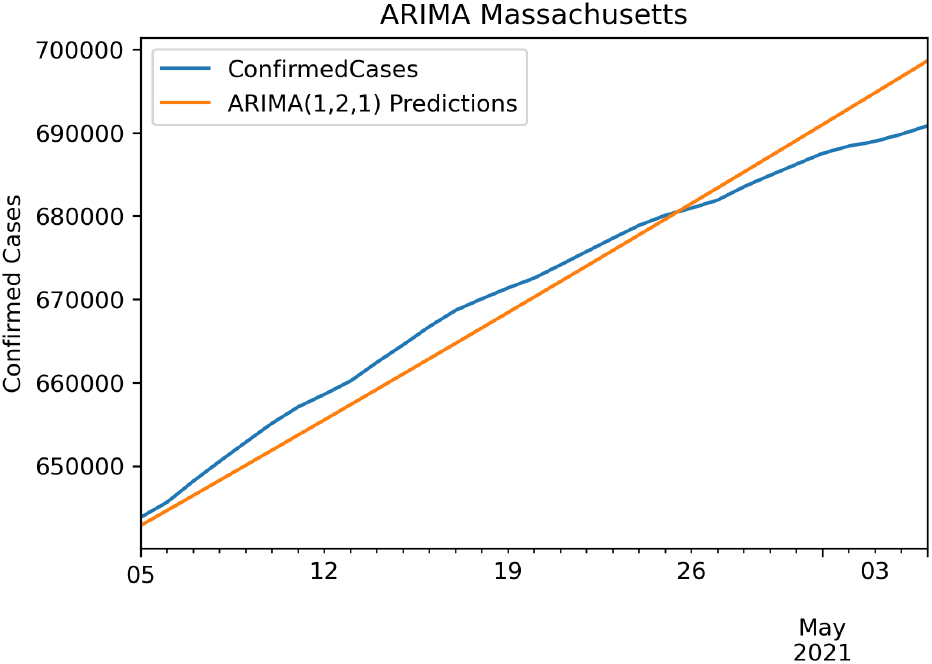
Massachusetts Confirmed ARIMA(1,2,1) RMSE: 2970.754175

**Fig. 28.**
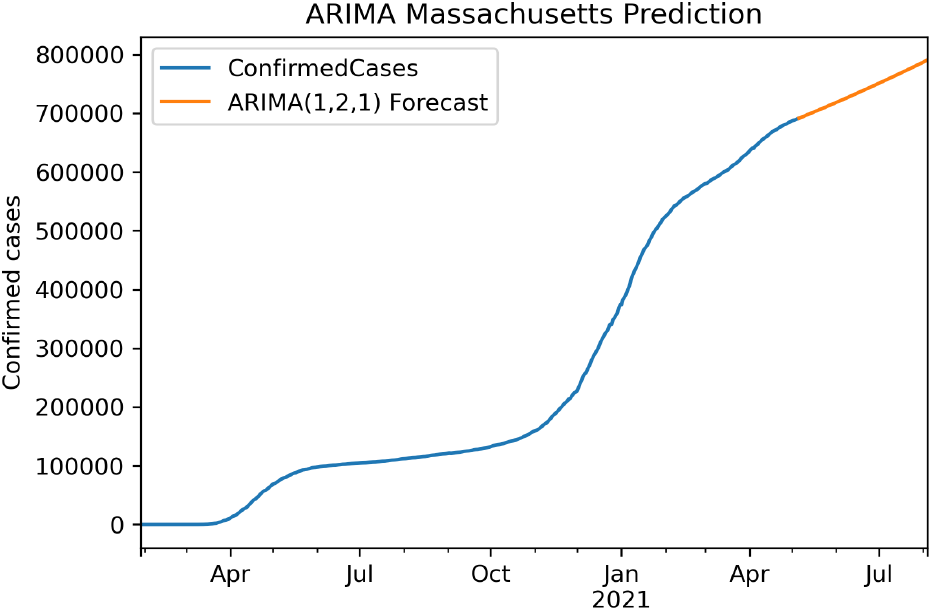
Massachusetts ARIMA Prediction

**Fig. 29.**
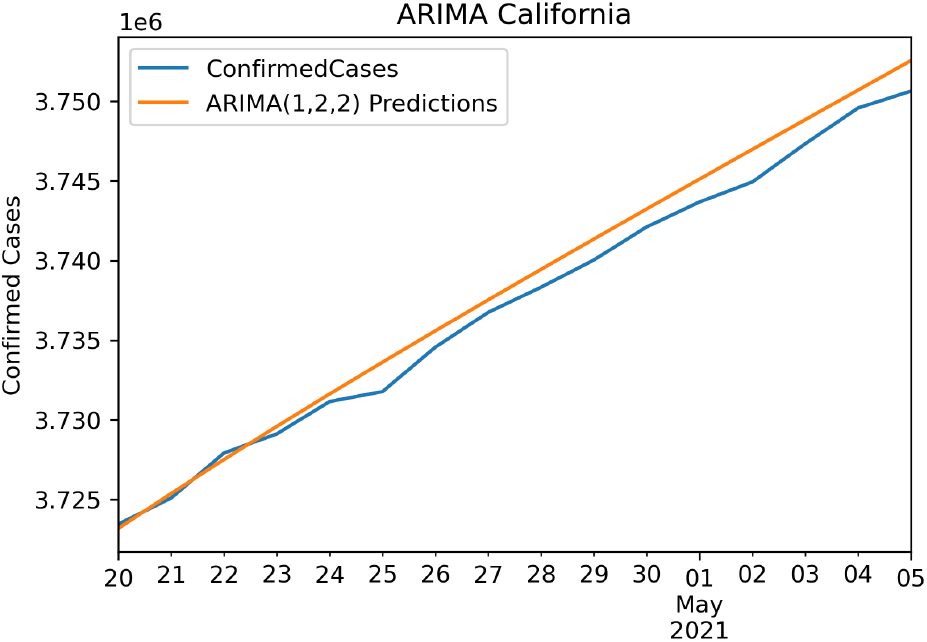
California Confirmed ARIMA(1,2,2) RMSE: 1012.258874

**Fig. 30.**
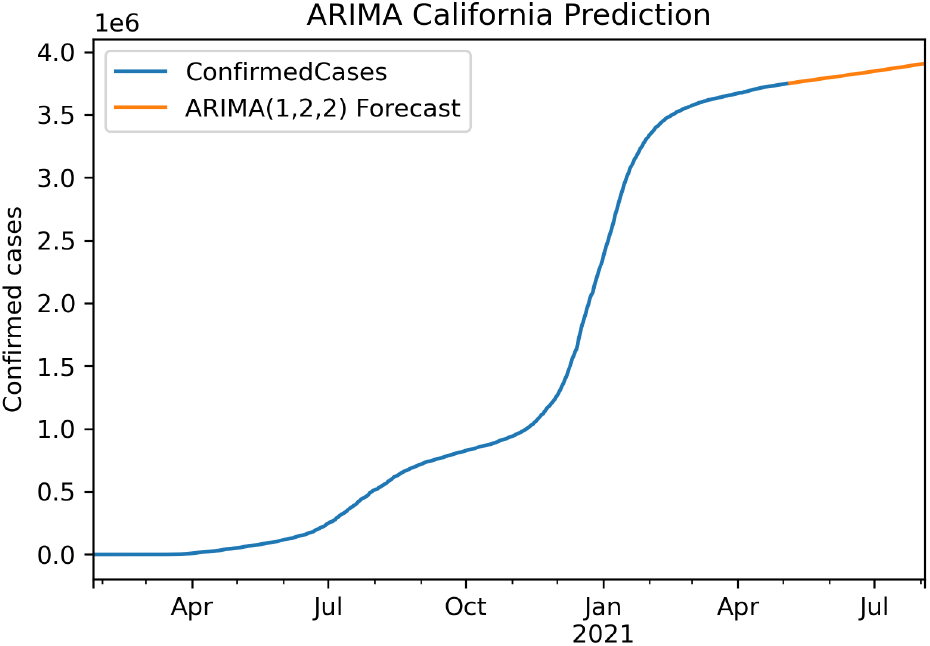
California ARIMA Prediction

**Fig. 31.**
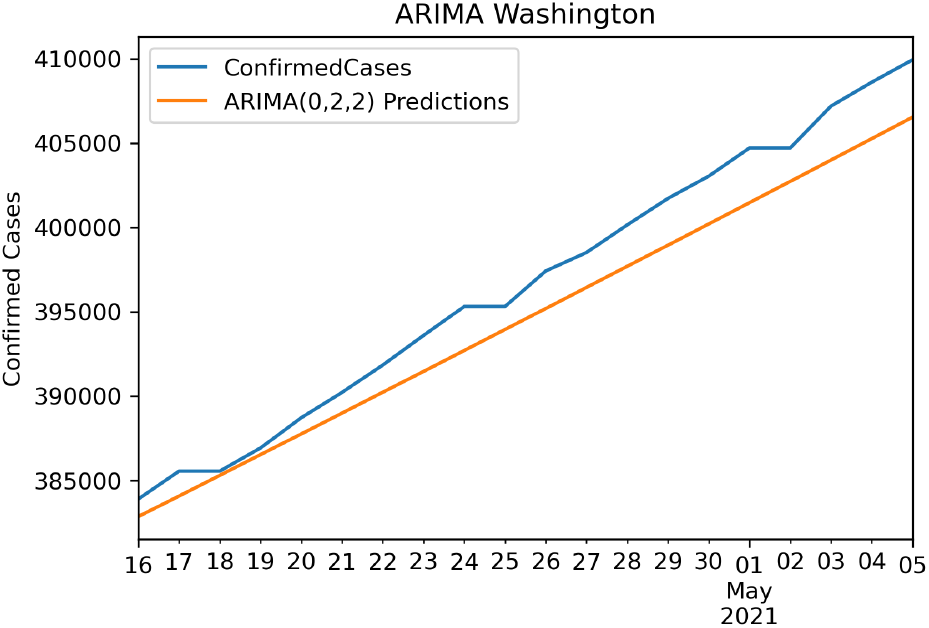
Washington Confirmed ARIMA(0,2,2) RMSE: 2238.090899

**Fig. 32.**
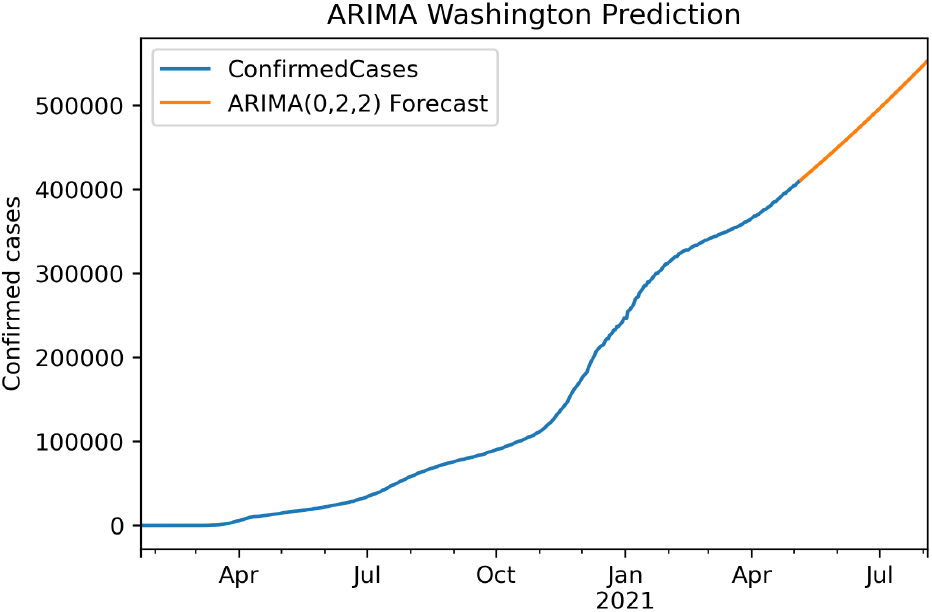
Washington ARIMA Prediction

**Fig. 33.**
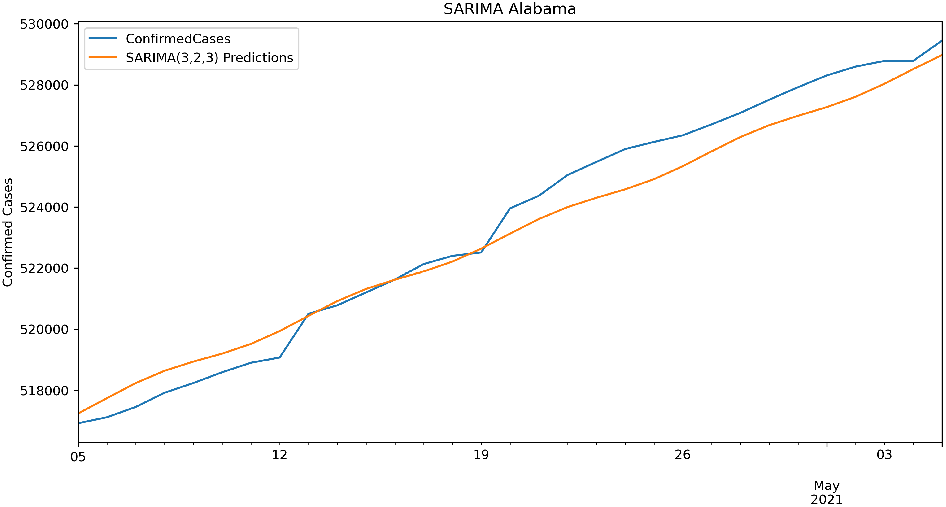
Alabama SARIMA(3, 2, 3)(2, 0, [1, 2], 2) RMSE: 754.7386732

**Fig. 34.**
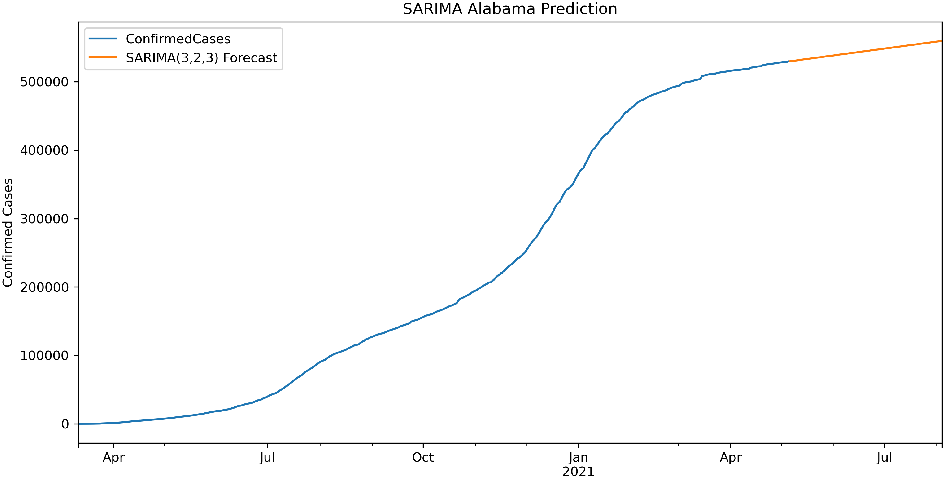
Alabama SARIMA Prediction

**Fig. 35.**
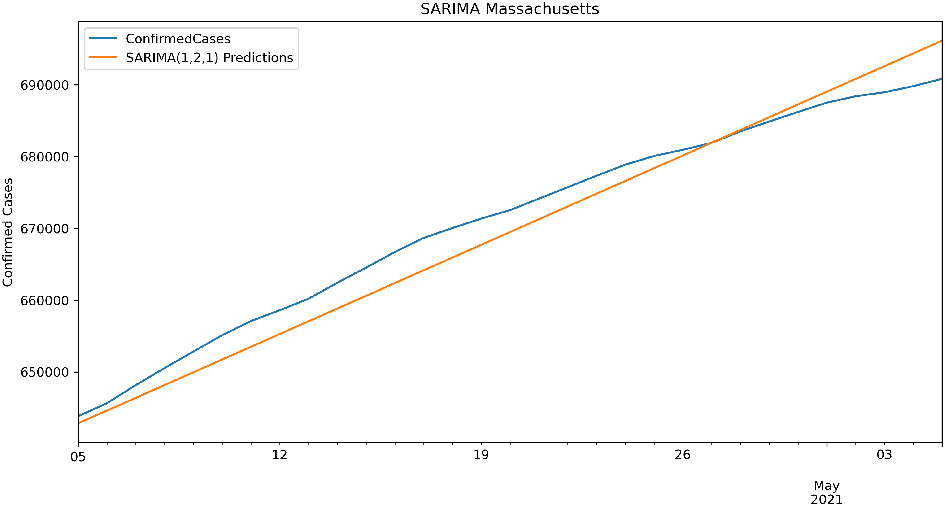
Massachusetts SARIMA(1, 2, 1)(2, 0, [], 2) RMSE: 3312.279949

**Fig. 36.**
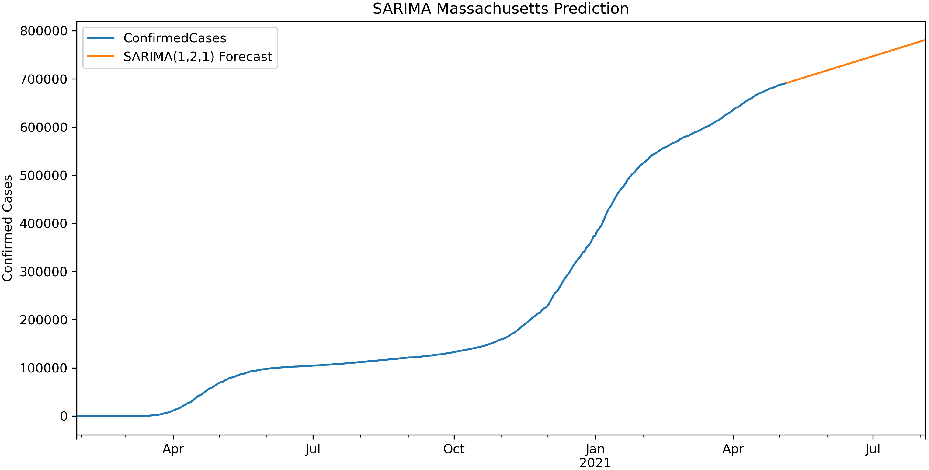
Massachusetts SARIMA Prediction

**Fig. 37.**
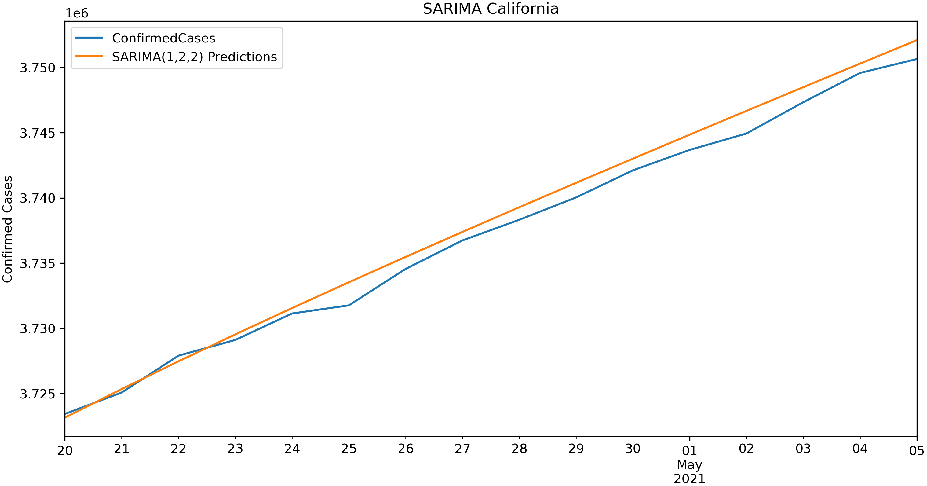
California SARIMA(1, 2, 2)(2, 0, 1, 2) RMSE: 1213.763069

**Fig. 38.**
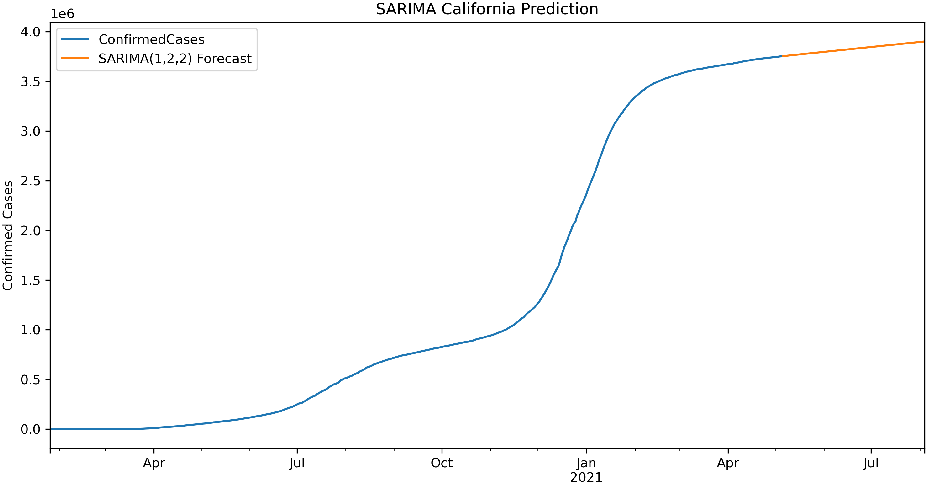
California SARIMA Prediction

**Fig. 39.**
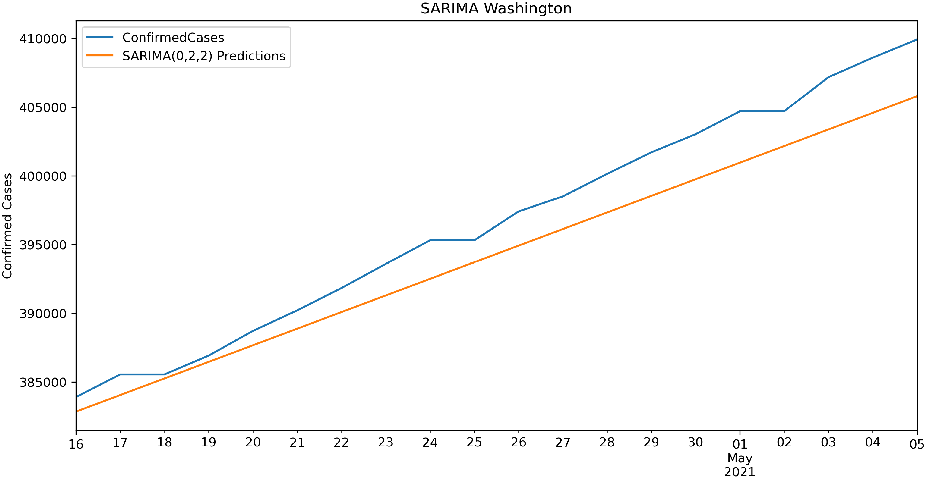
Washington SARIMA(0, 2, 2)(2, 0, 1, 2) RMSE: 2588.079917

## Conclusion

Time-series techniques have been used for a while to model the data, including data associated with time. This paper has applied three models of Simple Moving Average, Exponentially Weighted Moving Average, and Holt-Winters Double Exponential Smoothing Additive to model the number of infected individuals and the number of deceased COVID-19 cases. We have also used the ARIMA and SARIMA techniques to model and predict the number of cases. These models could help governors and healthcare providers to manage, plan and prepare for the peaks of this and similar diseases. The results were discussed using RMSE and MSE errors to be able to compare the performance of the methods to model and predict the number of cases. We have noticed that in the case of the three presented models, the Holt-Winters Double Exponential Smoothing Additive model outperforms the other two in both cases of the infected individuals and the deceased cases. Also, MSE shows that ARIMA models could be a better forecaster for the number of infected individuals rather than SARIMA. As the research limitation, we can mention the limited number of observations in the data set, which only covers 16 months. As it pertains to improving forecasting accuracy, the deep learning method could be applied to this data set.

**Fig. 40.**
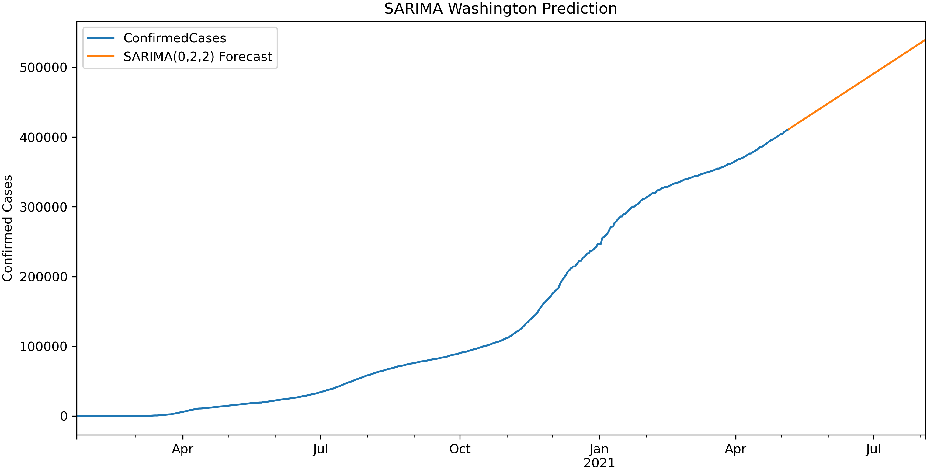
Washington SARIMA Prediction

## Data Availability

We are using the John Hopkins data repository

https://coronavirus.jhu.edu/map.html

## Authors’ contributions

**Saina Abolmaali**: Conceptualization, Methodology, Visualization, Formal Analysis, Writing-Original Draft **Samira Shirzaei**: Conceptualization, Visualization, Writing-Original Draft, Writing-Review and Editing.

